# Diastolic function and its relationship with the ATP-III criteria for metabolic syndrome in a Latin American cohort

**DOI:** 10.1101/2023.10.11.23296875

**Authors:** María Juliana Estévez Gómez, Diego Andrey Acevedo Peña, Silvia Fernanda Castillo Goyeneche, Anderson Felipe Arias, Luis Andres Dulcey Sarmiento, Jaime Gomez, Carlos Hernandez, Juan Sebastián Theran Leon, Raimondo Caltagirone, Edgar Blanco, María Ciliberti, Silvia Yulieth Calderon Amaya, Emily Gutierrez, Angie Lizcano, Maria Camila Amaya, Diego Lobo

## Abstract

**INTRODUCTION:** In South America, heart failure poses a substantial concern due to its widespread occurrence and insufficient data accuracy. Metabolic syndrome, a risk factor for heart disease and diabetes, is the focal point of the study, which seeks to examine its correlation with left ventricular diastolic function (LVDF) and indications of heart failure in adults within a South American hospital.

**MATERIALS AND METHODS:** A cross-sectional study with 2380 adults aged 65-79 in a South American hospital reveals connections between metabolic syndrome (MetS) and heart health. Using ATP-III criteria, MetS was identified, and cardiac function was assessed by echocardiography. Significant associations between MetS and various cardiac indicators were found.

**RESULTS:** Metabolic Syndrome (MetS) affected 33.1% of the sample, showing health differences and cardiac alterations, including ejection fraction changes. Associations with diastolic dysfunction criteria and complex relationships between natriuretic peptides and ventricular filling pressure were observed.

**CONCLUSIONS:** Metabolic syndrome links to significant changes in diastolic function and left ventricular structure, but not with alterations in the left atrium. Nevertheless, individuals with Metabolic Syndrome are more prone to receiving a diagnosis of chronic heart failure. Further extensive studies in diverse populations are advised.

## Introduction

Heart failure (HF) contributes significantly to premature disability and mortality [1]. Despite the high and growing prevalence of heart failure in South America, there is a lack of precise data on the prevalence of the disease at a regional level, which underlines the importance of epidemiological studies. The set of risk factors for the development of cardiovascular diseases (CVD) and type 2 diabetes mellitus (DM) associated with insulin resistance is known as metabolic syndrome (MS) [2]. The literature discusses the multicomponent mechanisms for the development of HF in SM, including the activation of microvascular endothelial inflammation and myocardial hypoxia with the development of coronary heart disease (CAD) and subsequent myocardial remodeling [3]. Recent analyzes provide insight into the fundamental basis of left ventricular (LV) dysfunction in heart failure with preserved ejection fraction (HFpEF), including systemic inflammation, coronary microcirculation abnormalities, cardiomyocyte stiffness, and myocardial fibrosis [4].

Diagnosis of heart failure includes collection of anamnestic data (including symptoms), evaluation of echocardiographic parameters (EchoCG) and determination of the level of natriuretic peptides in the blood. With preserved LV systolic function, to evaluate structural and borderline functional changes in the myocardium in obesity and other conditions associated with insulin resistance, the need to evaluate LV diastolic function (LVDF) increases. Despite the presence in clinical guidelines for the diagnosis of chronic heart failure (CHF) of a clear algorithm to identify diastolic dysfunction [5], to date, an extremely small number of population-based studies using this algorithm have been conducted.. The objective of the study was to evaluate the associations of metabolic syndrome with LVFD parameters and signs of heart failure in the adult population of a South American hospital.

## Material and methods

A cross-sectional study was carried out using a sample of the population of a South American hospital (n = 2380) between 65 and 79 years old, formed as part of the “Know” study. your Heart “(USS) in 2015-2017. Details of the sampling and data collection methods used in the study are published [6]. Those who accepted were interviewed in the hospital and medically examined using the ATP-III criteria for metabolic syndrome. To achieve the objectives of this study, an analysis of medical history data was carried out, including age, sex, smoking (never/past/current), presence of CHF, arterial hypertension (AH), coronary arteries, myocardial infarction, atrial fibrillation (AF) and diabetes diagnosed by a doctor in management. Anthropometric data were used: height measurements, waist circumference (WC) and hip circumference (CCa), calculation of body mass index (BMI) using the Quetelet formula. The analysis included measurements of systolic and diastolic blood pressure (SBP and DBP) (OMRON 705 IT automatic blood pressure monitor; (OMRON Healthcare Co., Ltd.). SBP and DBP levels They were measured three times with an interval of 2 minutes between measurements. The average values of the 2nd and 3rd measurements were used in the analysis. The following indicators were selected from the USS laboratory research database: total cholesterol (TC), high-density lipoprotein cholesterol (HDL), low-density lipoprotein cholesterol (LDL), triglycerides (TG), glycosylated hemoglobin (HbA1c), cystatin C, serum creatinine, high-sensitivity troponin T (hsTnT), N-terminal propeptide of brain natriuretic peptide (NT-proBNP), and blood high-sensitivity C-reactive protein (hsCRP).

transthoracic echocardiography were used, performed in one-dimensional and two-dimensional Doppler modes using a 1.5-3.6 MHz phased array sensor (Vivid q, (GE Medical Systems (China) Co., Ltd ., China)) in accordance with international recommendations [7].

Based on the results of echocardiography, the following indicators were evaluated:

— Left ventricular myocardial mass (LVM) indexed to body surface area.
— LV ejection fraction (EF) according to the Simpson method.
— LVFD parameters determined from transmitral flow and mitral annulus tissue Doppler data, including:
  a. E/A ratio (the ratio of the peak blood flow velocity from LV relaxation in early diastole to the peak blood flow velocity in late diastole caused by atrial contraction);
  b. (Early diastolic velocity of movement of the lateral part of the mitral annulus);
  c. (Velocity of the septal part);
  d. average E/e′ ratio. (early filling rate according to Doppler ultrasound transmitral /early relaxation rate according to tissue Doppler ultrasound);
  e. maximum and minimum volumes of the left atrium (OLP max and OLP min) ;
  e maximum and minimum indexed volumes of the left atrium (IOP max and IOP min).

When evaluating major and minor criteria for diastolic dysfunction, the consensus of international recommendations was used, in which the severity of changes is characterized as moderate deterioration (minor criterion) or severe deterioration (major criterion) (Table 1) [7, 8].

**Table 1.** Interpretation of left ventricular diastolic function and NT-proBNP levels.

| Index | Meaning | Interpretation |
| --- | --- | --- |
| **Myocardial functional indicators:** |  |  |
| **my'** | septal e' <7 cm/s<br>lateral e' <10 cm/s | major criterion |
| **Average e/e'** | >15 | major criterion |
|  | 9-14 | minor criterion |
| **EchoCG structural features:** |  |  |
| **CHEEP** | >34ml/ m<sup>2</sup> | major criterion |
|  | 29-34ml/ m<sup>2</sup> | minor criterion |
| **MVI** | ≥149 g/m<sup>2</sup> for men and ≥122 g/m<sup>2</sup> for women | major criterion |
|  | ≥115 g/m<sup>2</sup> for men and ≥95 g/m<sup>2</sup> for women | minor criterion |
| **Natriuretic peptides:** |  |  |
| **NT-proBNP level** | >220 pg/ml | Significant increase |
|  | 125—220 pg/ml | Moderate increase or |

Notes IOP: indexed left atrial volume; LVMI is left ventricular myocardial mass indexed to body surface area.

LVEF was considered reduced <40%; moderately reduced 40-49%; preserved >50%. According to the criteria proposed by the AHA/NHBLI (American Heart Association / National Heart, Lung, and Blood Institute) 2009 [2], SM was defined as the presence of three of the following five criteria: 1) WC ≥94 cm in men and ≥80 cm among women; 2) triglycerides (TG) ≥1.7 mmol /l; 3) HDL cholesterol <1.0 mmol /l for men and <1.3 mmol /l for women; 4) systolic blood pressure >130 and/or diastolic blood pressure >85 mm Hg. Art.; 5) fasting blood glucose level ≥100 mg/ dL (≥5.6 mmol /L). In this work, the level of glycosylated hemoglobin (HbA1c) >5.7% was used as an additional criterion. The present study included 2352 USS participants (982 men and 1370 women) who had the necessary data to determine the presence of metabolic work units (MetS).

### Statistical data analysis

Continuous variables are presented as means (M) with standard deviations (SD) or as medians (Me) with the first and third quartiles (Q1; Q3) depending on the distribution of the data. Comparisons between groups on continuous variables were performed using the independent samples t test. Continuous variables with skewed distributions were analyzed by transformation. Comparisons between groups in categorical variables were performed using the chi -square test (χ 2) Pearson test. The mean values of quantitative LVEF parameters in the groups with and without MS, as well as the proportions of participants with dichotomous signs of myocardial changes, were standardized by sex and age using the 2013 European standard population in the age range of 65 at 79 years old. years and are presented with 95% confidence intervals (DI). Associations of metabolic syndrome with quantitative indicators of LVEF were determined using multivariate linear regression adjusted for sex and age. The results of the linear regression analysis are presented as standardized β coefficients. The associations of MS with dichotomous signs (higher and lower LVEF criteria) were studied using multivariate logistic regression analysis adjusted for sex and age, the results of which are presented as odds ratio (OR) with 95% CI. SPSS 18.1 was used to perform statistical analyses.

### Ethical approval

The USS study was approved by the local ethical committees of the South American hospital where it was developed. All study participants provided signed informed voluntary consent.

## Results

The prevalence of MS in the analyzed sample was 33.1% (n = 778); among men - 32.8% (n = 322), among women - 33.3% (n = 456), p = 0.802. People with SM were on average 5.2 years older than participants without SM, but there were no differences by gender. In addition to differences in characteristics used to distinguish groups by the presence or absence of MetS, patients in the low MetS group were more likely to have a history of smoking, a history of CVD, and higher levels of biomarkers of inflammation (hsCRP) ., cystatin C), renal failure (creatinine), myocardial damage (high-sensitivity troponin T - hsTnT) and heart failure (NT-proBNP) (Table 2).

**Table 2.**
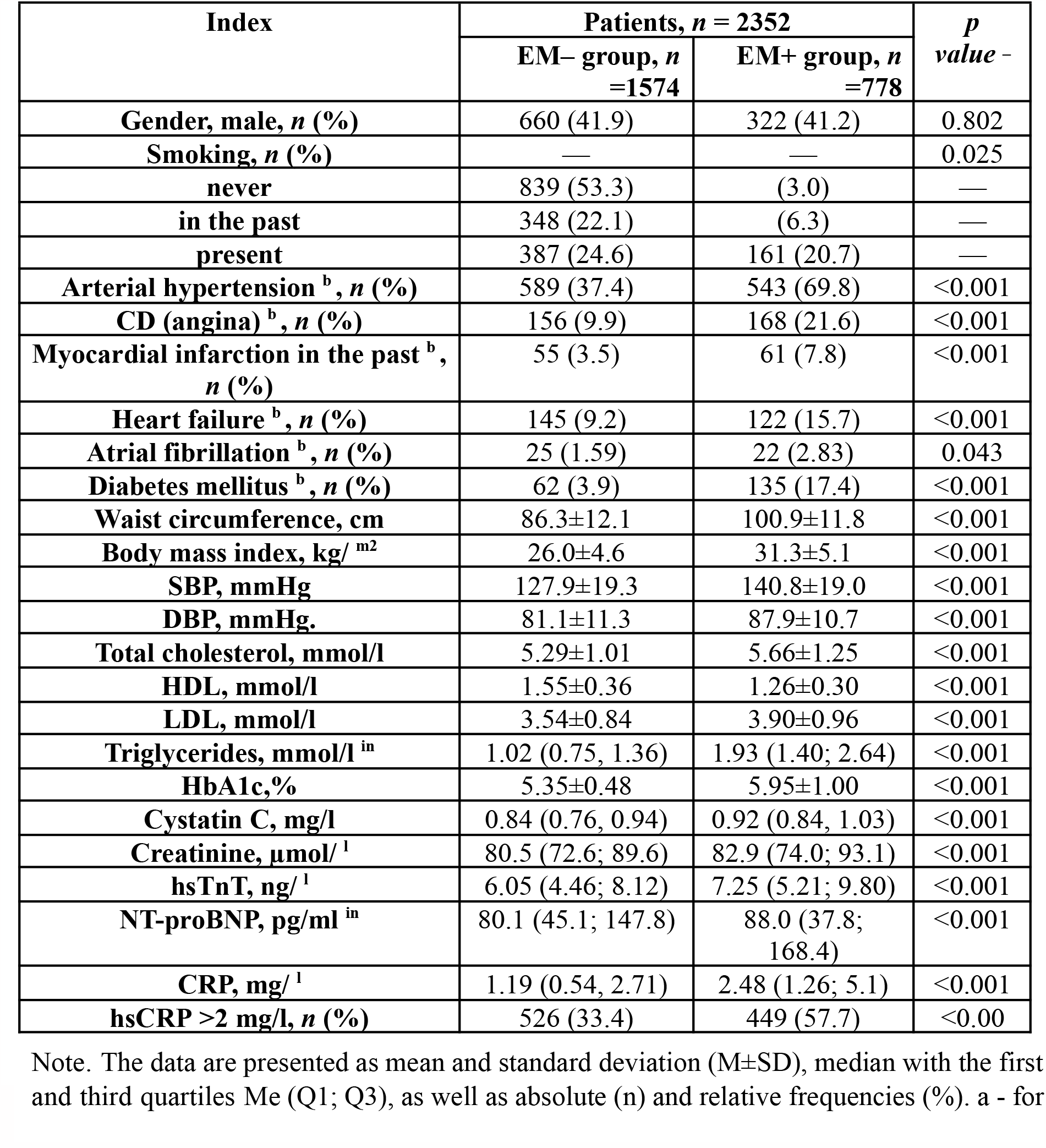
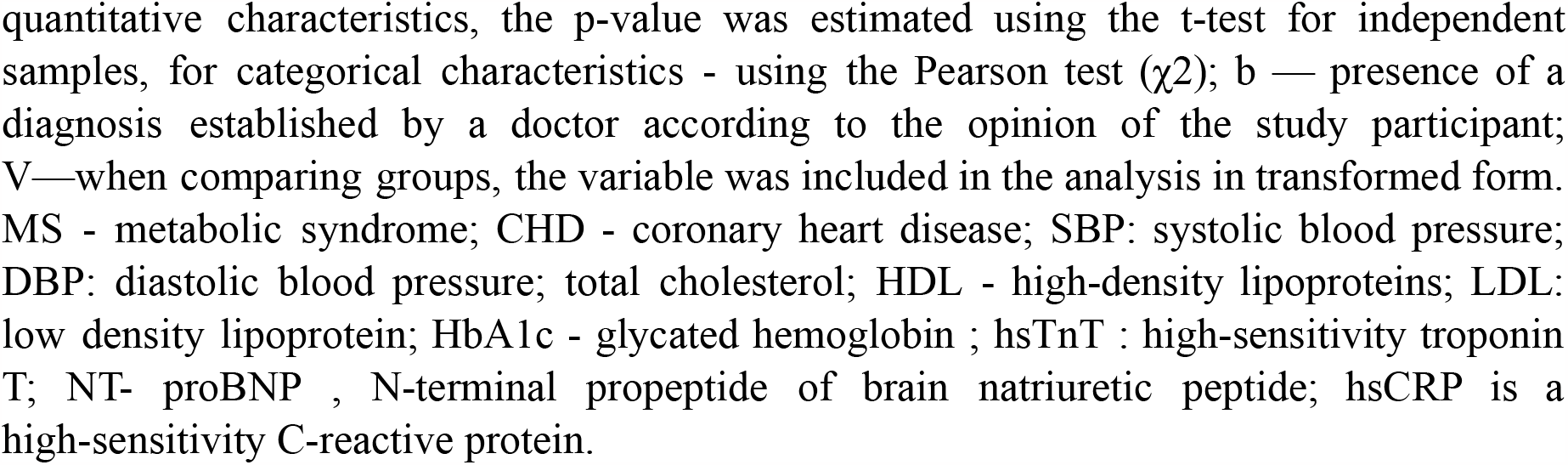
Characteristics of study participants with and without metabolic syndrome.

Reduced LV systolic function (EF <40%) was detected in 17 (0.8%) participants, including 12 (1.7%) participants in the MS group and only 5 (0.4%) in the MS group. without SM. Moderately reduced EF (40-49%) was detected in 198 (9.4%) participants, including 98 (14.0%) participants in the MS group and 100 (7.1%) in the non-MS group. The remaining participants had preserved EF (≥50%): 1,901 (89.8%) participants in the total sample, 588 (84.2%) in the MS group, and 1,313 (92.6%) in the non-MS group.. The distribution of participants in the groups with and without MS differed statistically significantly (p <0.001) between the EF categories.

The average EchoCG values of the DF indicators (lateral e′, septal e′, E/A ratio) standardized by sex and age were statistically significantly lower, and LV filling pressure (E/e′) was statistically significantly lower. higher in the group with MS compared to the group without MS (Table 3). The differences in the structural indicators of EchoCG were not so clear. Therefore, the standardized mean value of MVI was significantly higher in the individuals in the group with MS, as were the OLP max and OLP min indicators, but the OLP max and OLP min indicators did not have statistically significant differences.

**Table 3.** Average values standardized by sex and age* of functional and structural parameters of the myocardium in groups with and without metabolic syndrome and associations of indicators with metabolic syndrome.

| Index | Patients, $n = 2352$ | | Association of the indicator with MS <sup>a</sup> | |
| --- | --- | --- | --- | --- |
| | EM– group, $n = 1574$ | EM+ group, $n = 778$ | | |
| | $M_{\text{standard}}$ · (95% CI) | | Standardized coefficient $\beta$ | $P$ value - |
| Functional changes: |  |  |  |  |
| lateral e',<br>cm/s | 12.1 (12.0—12.2) | 10.6 (10.4—10.9) | –0.185 | <0.001 |
| septal e',<br>cm/s | 9.6 (9.5—9.7) | 8.3 (8.1—8.5) | –0.186 | <0.001 |
| E/e' | 7.0 (6.9—7.1) | 7.9 (7.7—8.1) | 0.167 | <0.001 |
| E/A ratio | 1.3 (1.3—1.3) | 1.1 (1.1—1.1) | −0.164 | <0.001 |
| Structural changes: |  |  |  |  |
| Max OLP, ml | 50.0 (49.3—50.8) | 54.1 (52.7—55.4) | 0.104 | <0.001 |
| Max IOP, ml/<br>m<sup>2</sup> | 27.1 (26.8—27.5) | 27.1 (26.5—27.7) | −0.010 | 0.644 |
| OLP<sub>min</sub>, ml | 21.8 (21.4—22.2) | 23.7 (22.9—24.5) | 0.081 | <0.001 |
| IOP<sub>min</sub>, ml/<br>m<sup>2</sup> | 11.8 (11.6—12.0) | 11.9 (11.5—12.2) | −0.001 | 0.952 |
| LVMI, g/m<sup>2</sup> | 109.4<br>(108.1—110.7) | 117.2<br>(114.7—119.7) | 0.114 | <0.001 |

Note. \* - direct standardization to the 2013 standard European population in the age range 35 to 69 years. a — results of the multivariate linear regression analysis with the corresponding indicator as the dependent variable and correction for sex and age. MS - metabolic syndrome; OLPmax and OLPmin : maximum and minimum volumes of the left atrium; IOLPmax and IOLPmin : indexed maximum and minimum volumes of the left atrium; LVMI is left ventricular myocardial mass indexed to body surface area.

When considering the relationships between SM and the major and minor criteria of diastolic dysfunction, the frequency of the criteria septal e <7 cm/s and lateral e <10 cm/s was high in the participants of both groups compared, while it was higher in the individuals in the group with SM (Table 4). The frequency of the minor criterion E/e′ (9-14) was also higher in individuals with MS. However, compliance with the major criterion (E/e′ ≥15) was found only in 9 (0.6%) and 11 (0.9%) participants in the groups without MS and with MS, respectively, which practically excluded the possibility of determining statistically significant differences due to the small number of cases.

**Table 4.** Sex- and age-standardized proportions* of participants with significant changes in structural and functional parameters of the myocardium, NT-proBNP and chronic heart failure in groups with and without metabolic syndrome and the association of these changes with metabolic syndrome.

| Indicators | Patients, *n* = 2352 |  | Association of the indicator with MS <sup>a</sup> |  |
| --- | --- | --- | --- | --- |
|  | EM group—, *n* =1574 | EM+ group, *n* =778 |  |  |
|  | Standard *P*. (95% CI), % |  | OR (95% CI) | *p* value - |
| Functional changes: |  |  |  |  |
| septal é <7 cm/s, % | 17.4<br>(15.7—19.2) | 32.6<br>(29.2-36.2) | 2.48 (1.99—3.08) | <0.001 |
| lateral é <10 cm/s,<br>% | 28.3<br>(26.4—30.4) | 42.6<br>(38.9—46.3) | 2.10 (1.71—2.57) | <0.001 |

| E/é, % |  |  |  |  |
| --- | --- | --- | --- | --- |
| <9 | 86.0<br>(84.3—87.6) | 75.1<br>(71.9—78.1) | Reference<br>category | — |
| 9-14 | 13.4<br>(11.8—15.1) | 23.9<br>(21.0—27.2) | 2.06 (1.64—2.58) | <0.001 |
| ≥15 | 0.6 (0.3—1.1) | 0.9 (0.5—1.8) | 1.75 (0.70-4.34) | 0.229 |
| Structural changes PIOP max, %: |  |  |  |  |
| <29 | 66.0<br>(63.6—68.3) | 67.0<br>(63.0—70.8) | Reference<br>category |  |
| 29—34 | 17.1<br>(15.3—19.1) | 17.9<br>(15.0—21.2) | 0.87 (0.71—1.08) | 0.209 |
| >34 | 16.9<br>(15.1—18.8) | 15.1<br>(12.4—18.3) | 0.96 (0.78—1.18) | 0.676 |
| in men | *n* = 660 | *n* = 332 | — | — |
| LVMI ≥115 g/m2 ·<br>% | 48.8<br>(45.1—52.6) | 58.3<br>(52.4—64.0) | 1.63 (1.23—2.16) | 0.001 |
| LVMI ≥149 g/m2 ·<br>% | 12.5<br>(10.2—15.1) | 17.0<br>(13.3—21.4) | 1.49 (1.03—2.14) | 0.032 |
| between women | *n* = 914 | *n* = 456 | — | — |
| LVMI ≥95 g/m2 · % | 56.3<br>(53.1—59.4) | 73.2<br>(67.2—78.5) | 1.84 (1.41—2.39) | <0.001 |
| LVMI ≥122 g/m2 ·<br>% | 17.1<br>(14.9—19.6) | 27.1<br>(22.0—32.9) | 1.45 (1.09—1.92) | 0.010 |
| Natriuretic peptides NT-proBNP pg/ml, %: |  |  |  |  |
| <125 | 70.4<br>(68.4—72.4) | 74.4<br>(71.1—77.3) | Reference<br>category | — |
| 125—220 | 17.0<br>(15.3—18.9) | 13.2<br>(11.0—15.8) | 0.76 (0.59—0.97) | 0.027 |
| >220 | 12.6<br>(11.1—14.2) | 12.4<br>(10.2—15.0) | 0.85 (0.65—1.12) | 0.249 |
| History of heart<br>failure<sup>b</sup>, % | 8.8 (7.5—10.2) | 11.4<br>(9.3—13.8) | 1.31 (1.00—1.72) | 0.04 8 |

Note. \* - direct standardization to the 2013 standard European population in the age range 65 to 79 years; Values for men and women are standardized by age. a — results of the multivariate logistic regression analysis with the corresponding indicator as the dependent variable and adjustment for sex and age (only for age in the LVMI analysis in the groups of men and women). b — presence of a diagnosis established by a doctor according to the study participant. MS - metabolic syndrome; IOLPmax : maximum indexed volume of the left atrium; LVMI, left ventricular myocardial mass indexed to body surface area; NT-proBNP , N-terminal propeptide of brain natriuretic peptide .

The frequencies of the major and minor criteria for IOP did not differ significantly in the groups with and without MS. Elevated MVI values (major and minor criteria) occurred with a statistically significantly higher frequency in individuals in the SM group, both men and women. In women, these differences were more pronounced, as were the associations of metabolic syndrome with the corresponding indicators.

The frequency of moderately elevated NT-proBNP values (125-220 pg /ml) was statistically significantly lower in individuals with MS, and the frequency of elevated NT-proBNP values (>220 pg /ml) had no statistical significance. significant differences. At the same time, study participants with MS more frequently reported a history of CHF (11.4%) compared to participants without MS (8.8%).

In individuals in the SM group, the indicator of moderate diastolic dysfunction (E/e′ in the range of 9-14) in 52.8% of cases was combined with a normal value of NT-proBNP (< 125 pg / ml), in 47.2% of cases - with an increase (≥125 pg /ml) (Table 5). A marked increase in the level of LV filling pressure (E/e′ >15) was accompanied by a moderate increase in the content of NT-proBNP (125-220 pg /ml) in 18.2% of cases., a significant increase (>220 pg /ml) /ml) in 72.7% of cases. The proportion of participants with a moderate increase in LV filling pressure (E/e′ in the range of 9 to 14) and normal NT-proBNP levels was 15.7% (n = 122) among all individuals with SM (n = 778).

**Table 5.**
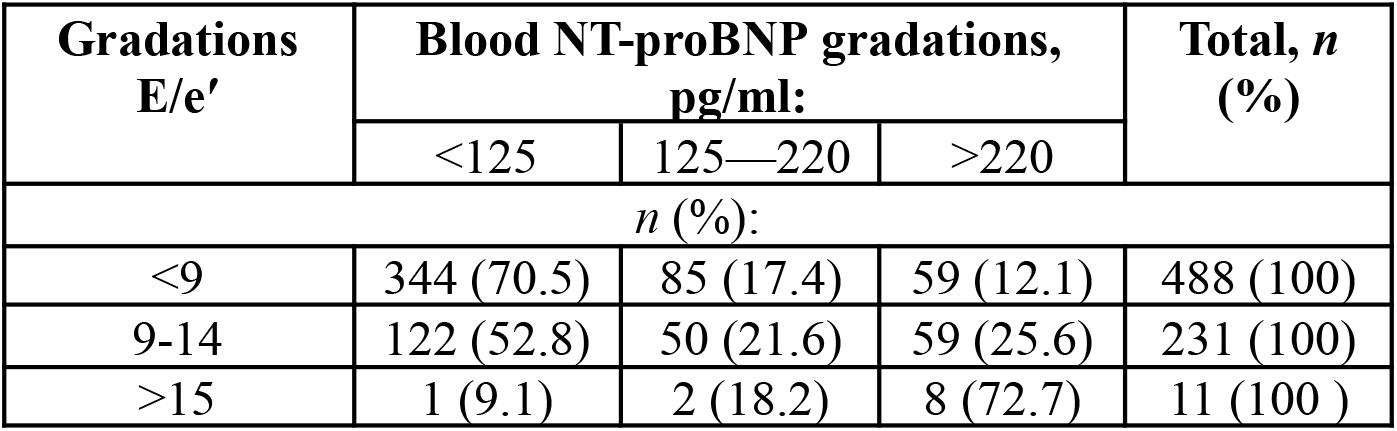
Association* of NT-proBNP with left ventricular filling pressure in patients in the metabolic syndrome group.

| Gradations<br>E/e' | Blood NT-proBNP gradations,<br>pg/ml: |  |  | Total, *n*<br>(%) |
| --- | --- | --- | --- | --- |
|  | <125 | 125—220 | >220 |  |
| *n* (%): |  |  |  |  |
| <9 | 344 (70.5) | 85 (17.4) | 59 (12.1) | 488 (100) |
| 9-14 | 122 (52.8) | 50 (21.6) | 59 (25.6) | 231 (100) |
| >15 | 1 (9.1) | 2 (18.2) | 8 (72.7) | 11 (100 ) |

## Discussion

SM was detected in 33.1% of a random sample of Arkhangelsk residents aged 35–69 years. When evaluating EchoCG parameters, preserved systolic myocardial function was revealed in the majority of participants (EF ≥ 50% was determined in 89.8%). Given that the majority of participants had a preserved or moderately reduced LVEF, the determination of the structural and functional disorders that characterize the diastolic function of the heart, as well as the levels of NT-proBNP, seems relevant to identify the risk of CHF. Since clinical symptoms and signs of heart failure were not analyzed in this article, heart failure was evaluated using anamnestic data.

When evaluating LVFD, a higher incidence of LV hypertrophy was observed in people with MS, and it was statistically higher in women. The frequency of moderate changes in E/e′ (9–14) was statistically significantly higher in individuals with SM. Furthermore, in 52.8% it was combined with a normal NT-proBNP value (<125 pg /ml) and in 47.2% with a high one. There were no differences in the frequency of changes in left atrial structure between the comparison groups. A similar result was obtained by other studies [9], which examined outpatients with preserved LVEF. Based on the mathematical models constructed, it was observed that the components of SM (DM, hypertension) are associated with changes mainly in the LV, but not in the LA. One of the most important risk factors for the development of diastolic dysfunction was age, along with type 2 diabetes and hypertension.

In our study, a history of CHF was associated with MS in the age group of 65 to 79 years. The sex- and age-standardized incidence of CHF in individuals in the group with MS (11.4%) was statistically significantly higher than in individuals without MS (8.8%). The literature discusses several main mechanisms for the development of heart failure in people with SM. First, obesity-related diseases and metabolic disorders are discussed as triggers leading to myocardial remodeling and diastolic dysfunction through microvascular endothelial inflammation [10]. In the sample of people with MS that we present, a high value of hsCRP, a marker of low-grade systemic inflammation, was detected in 57.7% of the participants, and this is statistically significant in people without MS. Previously, examining a sample of participants in another project, we also showed independent associations of hsCRP with both components of the metabolic syndrome and with functional and structural indicators of cardiac diastole [11]. Secondly, the development of cardiovascular risk factors associated with abdominal obesity has been described, leading to ischemic heart disease and myocardial infarction with subsequent formation of a fibrous scar and remodeling of the LV in the form of dilation and decreased systolic function., resulting in HF with reduced EF. [10]. This is consistent with our data on a higher frequency of reporting physician-diagnosed coronary heart disease and prior MI in the MS group than in the non-MS group and a higher frequency of low LVEF values (<40%). Third, the role of adipokines in the development of metabolic disorders and myocardial remodeling is suggested [12].

Thus, abdominal obesity and other components of SM are associated with various hemodynamic, neurohormonal and metabolic disorders that affect the structure and function of the heart [3]. In some studies based on the results of an analysis of clinical trial data, they described 3 phenotypes of patients with HF and preserved EF (HFpEF) and their response to a series of drugs used for treatment, in particular, spironolactone [13]. The comparison of the data we obtained on the nature of diastolic dysfunction in the MS group with the main characteristics of the proposed phenotypes allowed us to conclude that the study participants with MS are similar to the second phenotype (obesity, metabolic, systemic disorders)., a predominant moderate increase in the level of LV filling pressure (E/e’ value) and an increase in NT-proBNP only in 50% of individuals with an E/e’ value in the range of 9-14. To the literature, Obese patients with heart failure have lower concentrations of NT-proBNP due to increased expression of scavenger receptors and increased degradation of the peptides by adipose tissue [14]. It has previously been shown that people with HFpEF and normal NT-proBNP levels had a higher risk of death or rehospitalization from HF compared to people without HF [15]. Clinical studies have shown that patients with HFpEF and low levels of NT-proBNP responded better to mineralocorticoid receptor antagonists and angiotensin receptor blockers compared to patients with higher levels of NT-proBNP [16 -twenty]. The importance of identifying asymptomatic individuals with signs of moderate diastolic dysfunction and normal NT-proBNP levels is determined by the open prospect of preventing the development of severe diastolic dysfunction and symptomatic heart failure in them.

Our study has a number of limitations. First, a cross-sectional design was used, which limits the possibility of making statements about the cause-and-effect nature of the relationships identified between MS and LVEF and HF indicators. Secondly, the study carried out in a population sample of relatively healthy people between 65 and 79 years old did not allow us to identify a sufficient number of participants with severe deterioration of LVEF and signs of heart failure to determine their associations with MS.

Therefore, the study focuses more on the connections between SM and the initial changes in the structural and functional parameters of the myocardium, which, from the point of view of prevention, may be its strong point. Another limitation is that echocardiography did not evaluate the rate of tricuspid regurgitation, which would give an idea of the participation of the right ventricle in the formation of HFpEF and the presence of severe diastolic dysfunction.

## Conclusion

Study participants with metabolic syndrome had greater changes in diastolic function compared to those without metabolic syndrome. The frequency of changes in the structural parameters of the left ventricle was greater in the presence of metabolic syndrome. There were no associations of structural changes in the left atrium with metabolic syndrome. Functional diastole disorders were mainly characterized by a moderate increase in the filling pressure level of the left ventricle. In more than half of people with metabolic syndrome, a moderate increase in left ventricular filling pressure was not accompanied by an increase in NT-proBNP, consistent with current knowledge of lower NT-proBNP levels in overweight and obesity. However, study participants with metabolic syndrome were more likely to report having a medical diagnosis of chronic heart failure than participants without metabolic syndrome. Statistically significant associations of the metabolic syndrome with indicators of left ventricular diastolic function and anamnesis data on the presence of chronic heart failure were identified. Based on this, we recommend carrying out larger studies in other populations in our latitudes.

## Data Availability

All data produces in the present work are contained in the manuscript.

## Notes

### Competing Interest Statement

The authors declare no conflict of interest

### Funding Statement

This study did not receive any funding.

### Author Declarations

The ethics committee of the University of Los Andes reviewed this study in the year 2015 supporting its execution.

